# COVID-19 Pandemic Among Immigrant Latinx Farmworker and Non-farmworker Families: A Rural-Urban Comparison of Economic, Educational, Healthcare, and Immigration Concerns

**DOI:** 10.1101/2020.10.30.20223156

**Authors:** Sara A. Quandt, Natalie J. LaMonto, Dana C. Mora, Jennifer W. Talton, Paul J. Laurienti, Thomas A. Arcury

## Abstract

COVID-19 has highlighted social and health injustices in the US. Structural inequalities have increased the likelihood of immigrants contracting COVID-19, by being essential workers and through poverty that forces this population to continue working. Rural and urban immigrant families may face different concerns. Using a telephone survey in May 2020 of 105 Latinx families in an existing study, quantitative and qualitative data were gathered on work and household economics, childcare and education, healthcare, and community climate. Analyses show that, although rural and urban groups experienced substantial economic effects, impacts were more acute for urban families. Rural workers reported fewer workplace protective measures for COVID-19. For both groups, fear and worry, particularly about finances and children, dominated reports of their situations with numerous reports of experiencing stress and anxiety. The experience of the pandemic is interpreted as an example of contextual vulnerability of a population already experiencing structural violence through social injustice.

## Introduction

The COVID-19 pandemic has highlighted social and health injustices in the United States (US) population.^1^ Both case rates and mortality are higher among people of color than among white individuals.^2,3^ Immigrants of color and their families are among those groups at the highest risk for contracting COVID-19.^4,5^

Immigrants’ particular vulnerability to both acquiring the coronavirus and then developing severe COVID-19 appears to lie in structural inequalities.^6^ Many work in occupations and industries deemed essential, so that they have been exempted from protective stay-at-home or work-from-home orders.^7-9^ In addition, many immigrants live in crowded housing and multi-family dwellings that do not permit social distancing.^10^ These risk factors for exposure to COVID-19 are exacerbated by the fact that many immigrants have low incomes, cannot communicate well in English, and lack documentation status to be in the US and to access health and income safety net programs.^11,12^ For many immigrants, even if their particular job is not deemed essential, their low incomes and limited savings force them to continue working.^13^ Most have received no benefits from the special allocations in the Coronavirus Aid, Relief and Economic Security (CARES) Act distributed in spring and summer of 2020.^14^

The industry sectors where immigrants predominate and are deemed essential include farm work in agriculture and meat and poultry processing in manufacturing.^4,7,8,15-17^ As both of these industry sectors are crucial to the food system, there is considerable pressure for production to keep food availability at normal levels. Numerous outbreaks have been reported throughout the pandemic among these workers.^18-22^ These workers are particularly vulnerable because, in addition to not being able to work from home, their jobs usually prevent them from maintaining physical distancing in the workplace. Farm work, for example, often entails transportation in crowded vehicles to the fields and working side-by-side on equipment for planting or harvesting. Meat processing requires workers to be side-by-side on crowded production lines, without barriers. In all cases, communication is frequently by shouting or loud talking over machine noise, which is known to expel more virus-containing droplets or aerosols than speaking normally.^23^

Immigrant workers share the concerns of other US residents about the threats of the COVID-19 pandemic to their health and that of their families.^10^ However, their concerns may extend to anticipated outcomes in other aspects of life. Clark et al.^4^ suggest that anticipated outcomes may fall into three domains in addition to health and medical: economic (including fewer days or hours of work, job loss, and inability to pay bills including rent); legal (lack of eligibility for unemployment and other benefits), and social (including food insecurity, children’s difficulty with obtaining quality education, and worsening political and community climate surrounding immigration).

In research to date about fears in the pandemic among immigrant workers, health has been emphasized (e.g., ^10^). Less attention has been paid to concerns of immigrant workers in these other domains. In particular, few studies attempted to document these concerns during the early, confusing days of the pandemic and to do so in the words of these immigrants. In addition, there has been little comparison of the experiences of Latinx immigrants during the pandemic in rural and urban environments. It is likely that the experiences differ, based on differences between rural and urban situations in resources, in the demand for immigrant labor, and in the availability of public health messaging about the pandemic.

This paper reports findings from a telephone survey collected in May 2020. The respondents were participants in an existing study that had developed trust and lines of communication with mothers in a sample of rural Latinx farmworker families and a comparison group of urban Latinx non-farmworker families in North Carolina, USA. The survey gathered both quantitative and qualitative data from these mothers. In this paper, we describe the experiences of these workers’ families in four domains: work and household economics, childcare and education, healthcare, and community social climate concerning discrimination and racism. Throughout this description, we compare the experience of rural and urban immigrant Latinx families, and we use qualitative responses of the women to highlight the quantitative findings. We interpret our findings with a view to the social injustices these workers face as immigrants to the US.

## Methods

### Research design

The study reported here is part of a larger two-group, prospective study examining the health and cognitive effects of pesticide exposure in children in farmworker families. The larger study uses a comparative design, with a sample of families of Latinx farmworkers with children and a sample of similar families, but without any farmworker members. Additional details of the larger study can be found elsewhere.^24^ The current study used a telephone survey to reach the mother of the children in these families in May, 2020, when no face-to-face contact between study staff and study participants was permitted by the Institutional Review Board due to COVID-19-related health concerns for research participants. All procedures, including the telephone survey, were approved by the Wake Forest University Institutional Review Board. The study received a Certificate of Confidentiality from the National Institutes of Health.

### Inclusion criteria and participant recruitment

Inclusion criteria for the families were similar in both samples when recruited from March, 2018, to December, 2019; they reflect the purpose of the larger study. Each family had to have a child aged 8 years at baseline, who had completed the first grade in the US. All children had to be from families that self-identified as Latino or Hispanic, and with household incomes below 200% of the US federal poverty guideline. In the farmworker sample, the mother or her spouse must have been employed in farm work on non-organic farms during the past three years. In the non-farmworker sample, adults could not have been employed in any industry that involves routine exposure to pesticides (e.g., farm work, landscaping, pest control) in the previous three years. Families in the non-farmworker sample could not have lived adjacent to agricultural fields in the previous three years.

Exclusion criteria for both samples included children having life threatening illnesses, prior history of neurological conditions, physical condition or development disorder that would not allow them to complete or would interfere with the results of neurobehavioral tests or MRIs (used in the larger main study), primary language other than Spanish or English spoken in the home, or refusal of mother/guardian to complete the questionnaires.

In the larger study, a total of 76 children were recruited for the farmworker sample and 65 children for the non-farmworker sample. For the recruitment of the original sample, the North Carolina Farmworkers Project developed a list of farmworker families with an 8 year old child, and the locations where they lived. In addition, other community organizations that served farmworker families in the recruitment area were contacted. Study personnel contacted the mothers. Similarly, for the original non-farmworker sample, local recruiters in Winston-Salem, NC, and community members developed a list. For both samples, mothers were contacted by a bilingual staff member who explained the overall study procedures, answered questions, and, if the mother agreed to participate, obtained signed informed consent from the mother and assent from the child. As recruitment progressed, community partners worked with the study team to balance the two samples on gender of the child.

Prior to the telephone survey, 5 children in the farmworker sample and 17 in the non-farmworker sample withdrew, moved away from the study area, or were lost to follow-up. The remaining children represented 67 farmworker families and 45 non-farmworker families, because some families had more than one child enrolled. For the telephone sample, 2 families refused to participate and 5 could not be reached, all in the non-farmworker sample. A total of 67 farmworker families and 38 non-farmworker families could be reached and agreed to participate. This sample of 105 is used in this paper.

### Data collection

Details of data collection can be found elsewhere.^10^ Briefly, data for this study were gathered from May 1, 2020, to June 5, 2020, using a telephone survey. Only two interviews were conducted in June. Interviewers were members of the larger study team who had usual interview contact with the mothers. Interviewers received individualized televideo training and completed practice interviews. To recruit participants, interviewers called the last known telephone number for the mother in each family, explained the purpose and procedures for the study, and told the mother that she would receive a $10 incentive for completing it. The interviewers attempted calls at different times of day until the participant was reached or at least 3 unsuccessful calls had been made.

If the mother agreed to participate, her informed consent was noted, and the interviewer proceeded to conduct a standardized interviewer-administered questionnaire in the language of her choice. Data were entered in real time during the interviews using Research Electronic Data Capture (REDCap). The REDCap system provides secure, web-based applications for a variety of types of research.^25^ Data from this interview were later merged with selected personal, family, and household variables collected in the main study questionnaires.

### Variables and measures

Variables from the main study baseline questionnaire were used to create measures to describe the sample. These included the following measures for the mother: age, preferred language, country of origin, educational attainment for mother and spouse, and assignment of the family to the farm work or non-farm work sample.

#### Work and household economics

Employment status of the mother was obtained by asking what statement best reflected her current employment status relative to her primary job before the pandemic. Response options were: did not have a primary job before the pandemic, still working in the same job as before the pandemic, working in a different job from that at the start of the pandemic, lost her primary job and was looking for work, temporarily laid off from primary job, or on sick leave or other leave from primary job. These were collapsed to create a measure of current work status which included the category no primary job before pandemic (not in the labor force), same job as before pandemic, working a different job than before the pandemic, and currently out of work (laid off, on leave).

Participants were also asked about current work compared to the start of the pandemic. Response options were that hours were fewer than usual, more than usual, or about the same. To gauge protections from the coronavirus at work, participants were asked which of a series of protective measures her employer had implemented: provided masks, gloves, other personal protective equipment (PPE), and hand sanitizer; had instituted more frequent cleaning of workplace floor and other surfaces than usual; had erected partitions between workers; had moved desks or workstations farther apart; established daily health checks; and closed break or eating spaces. The same questions were asked for the spouse’s employment and workplace situation.

Open-ended questions were asked to ascertain whether there were any specific circumstances at the participant’s or her spouse’s work that made them worry about the coronavirus. Responses were recorded verbatim.

To measure expected economic status, participants were asked what the chance was that their family would run out of money in the next three months. Response options were very likely, somewhat likely, possible, unlikely, and very unlikely.

Three questions ordered from least to most severe were used to obtain a measure of household food security. These questions were based on a Spanish-language adaptation^26^ of the US Household Food Security Survey Module.^27^ Each specified “in the last seven days” and behavior “because of a lack of money or other resources”. These were (1) were you worried you would run out of food? (2) did anyone in the family eat less than they thought they should? (3) did anyone in the family go without eating for a whole day? A food security score was calculated based on the number of statements affirmed, ranging from 0 (food secure) to 3 (lowest food security).

Six situations unrelated to money that made it difficult to obtain food were queried: grocery stores open fewer hours, grocery stores make you wait in line to enter the store, grocery stores do not have the food you want, children are not getting school breakfast or lunch, or restaurants where you usually eat are closed. Those not affirming any of these options were classified as having no problems getting food.

#### Childcare and education

To assess how families accommodated school closures, mothers were asked how their children were being cared for when they were at work. Response options were: spouses work at different times (or one or both parents do not work), leave children at home alone (including having an older sibling care for young siblings), friend or relative provides childcare, or children attend licensed daycare center.

Two questions were used to obtain information on how education was provided while schools were closed. These questions were (1) did any of your children receive books or assignments from their teachers when they were sent home? and (2) have any of your children been receiving instruction over the internet? For both questions, response options were yes or no.

Participants were also asked to list their concerns about their child’s education while they were out of school. Responses were coded into the following options: children will fall below their grade level, children will not graduate or be promoted to the next grade, children will do poorly on the state end-of-grade tests, children can not take the ACT, SAT, or other college tests. If the mother had no concerns, this was noted. For additional information on mothers’ concerns about school closures, participants were asked to respond to three statements using a 5-point Likert-type scale: (1) school closures have made it difficult for me to work or do other household tasks, (2) I am satisfied with the communication from my children’s schools to support learning, and (3) my children will be prepared for school in the next school year. Response options were strongly disagree, disagree, neutral, agree, or strongly agree. The first two and last two response options were collapsed to create a measure with three response options: disagree, neutral, agree.

#### Healthcare

To gauge how the pandemic has altered families’ use of healthcare services, the participants were asked a series of three questions that looked at their actions within the past two weeks. Response options for all questions were no or yes. The first question asked if they or a or a family member had visited a doctor or clinic. The second question asked participants if they or a family member had canceled a planned doctor or clinic visit. If participants replied yes, they were asked two follow up questions: (1) did you cancel it because of the coronavirus?, and (2) did you cancel it because of the cost or lack of insurance? If participants answered yes to either question, they were asked to explain why. The third question asked participants if they or a family member had made any other change in medical care due to the coronavirus.

#### Community attitudes

Two questions were asked to assess current community attitudes on political and social issues: (1) do you think community members are more worried than usual about discrimination and racism?, and (2) do you think community members are more worried than usual about immigration issues? Response options for both questions were no, yes, and don’t know. If participants answered yes, they were asked to explain their answer and their responses were recorded verbatim.

To obtain measures on mother’s concerns during the COVID-19 pandemic, outside of the ones addressed in the questionnaire, participants were asked an open-ended question: is there anything else you want to tell us about your situation now in the pandemic? Responses were recorded verbatim.

### Data analysis

Frequencies and percents were calculated to examine the variables of interest by farmworker status and significant differences were examined using Chi-Square or Fisher’s Exact tests as appropriate. All analyses were done using SAS v 9.4 (SAS Institute, Cary, NC) and p-values < .05 are considered statistically significant.

Answers to open-ended questions about employment and economic concerns, about community attitudes, and about overall concerns during the pandemic were translated to English. These were then reviewed by the authors and themes identified. A list of codes was developed that included both themes and attitudes or emotions conveyed by the respondents through words used. These codes were applied to the text segments. Those codes that occurred across a number of participants were considered salient^28,29^ and were summarized for presentation. Exemplary quotations were chosen for presentation.

## Results

### Description of the sample

The women interviewed ranged from 25 to 47 years of age (Table 1). Most in both samples were born in Mexico; Spanish was the preferred language for almost all women. Years of formal education completed by the respondents ranged from no formal education to college graduate, with a median in both samples of ninth grade. Formal education of spouses was slightly less, with medians of sixth grade for the farmworker sample and eighth grade for the non-farmworker sample.

**Table 1.** Individual and household characteristics of participants. Comparison of Latinx farmworker and nonfarmworker adults in North Carolina, May 2020.

|  | Farmworker<br>n=67 |  | Non-Farmworker<br>n=38 |  |
| --- | --- | --- | --- | --- |
|  | n | % | n | % |
| Age |  |  |  |  |
| 25 – 29 years | 7 | 10.45 | 5 | 13.16 |
| 30 – 34 years | 26 | 38.81 | 7 | 18.42 |
| 35 – 39 years | 19 | 28.36 | 13 | 34.21 |
| 40 – 47 years | 15 | 22.39 | 13 | 34.21 |
| Country of birth (mother) |  |  |  |  |
| Mexico | 54 | 80.60 | 30 | 78.95 |
| El Salvador | 7 | 10.45 | 0 | 0 |
| Guatemala | 2 | 2.99 | 1 | 2.63 |
| Honduras | 1 | 1.49 | 3 | 7.89 |
| United States | 3 | 4.48 | 2 | 5.26 |
| Other | 0 | 0 | 2 | 5.26 |
| Language most comfortable for conversation |  |  |  |  |
| Spanish | 65 | 97.01 | 35 | 92.11 |
| English | 1 | 1.49 | 3 | 7.89 |
| An indigenous language | 1 | 1.49 | 0 | 0 |
| Highest level of education completed (mother) |  |  |  |  |
| Less than sixth grade | 13 | 19.40 | 3 | 7.89 |
| Sixth - eighth grade | 18 | 26.87 | 8 | 21.05 |
| Ninth – eleventh grade | 25 | 37.31 | 16 | 42.11 |
| High school or more | 11 | 16.42 | 11 | 28.95 |
| Highest level of education completed (spouse)<sup>1</sup> |  |  |  |  |
| Less than sixth grade | 13 | 23.64 | 7 | 19.44 |
| Sixth - eighth grade | 17 | 30.91 | 11 | 30.56 |
| Ninth – eleventh grade | 20 | 36.36 | 7 | 19.44 |
| High school or more | 5 | 9.09 | 11 | 30.56 |
| Household composition |  |  |  |  |
| Number of children |  |  |  |  |
| 0-1 | 4 | 5.97 | 1 | 2.63 |
| 2-3 | 40 | 53.70 | 22 | 57.89 |
| 4 or more | 23 | 34.33 | 15 | 39.47 |
| Number of adults |  |  |  |  |
| 1 | 7 | 10.45 | 2 | 5.26 |
| 2 | 45 | 67.16 | 24 | 63.16 |
| 3 or more | 15 | 22.39 | 12 | 31.58 |
| Total household size |  |  |  |  |
| 1-3 | 6 | 8.96 | 1 | 2.63 |
| 4-6 | 49 | 73.13 | 25 | 65.79 |
| 7-13 | 12 | 17.91 | 12 | 31.58 |

<sup>1</sup> Totals 55 and 36, respectively, due to missing values.

Medians for total household size were 5 (range 1 to 10) and 6 (range 3 to 13) in the farmworker and non-farmworker samples, respectively. The number of adults in the household for the farmworker sample ranged from 1 to 6, while the number of children ranged from 0 (for a respondent currently separated from her family) to 7. For the non-farmworker sample, the ranges were 1 to 4 for adults and 1 to 10 for children.

At baseline (March 2018 through December 2019), farmworker families reported that the most common industry in which women worked was agriculture; for men, it was construction, followed by agriculture. For non-farmworker families, most women were not in the labor force and the majority of men worked in construction.

### Comparison of Household Concerns

#### Work

About 40% of the women in both samples did not work in a regular job before the pandemic (Table 2). The women in farmworker families appeared to have less job stability: fewer were working in the same job at the time of the interview, and more were in a different job. About the same proportion (about 16%) of both samples of women reported being out of work. Among spouses in both samples, most were in the same primary job as before the pandemic. About 5% were in a different job, and only a few were out of work. Overall, the pandemic appears to have affected the employment of women in farmworker families more than any other category. Among those women, only 13 (34.21%) of the 38 who reported being in the labor force were working in the same job as prior to the pandemic, compared to 91.53% of their spouses. However, 59.09% and 86.11% of the women and spouses, respectively, in non-farmworker families, were in the same job.

**Table 2.** Comparison of employment status and COVID-19 accommodations for respondent and spouse in farmworker and non-farmworker families.

|  | Self |  | Spouse<sup>1</sup> |  |
| --- | --- | --- | --- | --- |
|  | Farmworker<br>n (%) | Non-Farmworker<br>n (%) | Farmworker<br>n (%) | Non-Farmworker<br>n (%) |
| Current employment status |  |  |  |  |
| No primary job before pandemic | 29 (43.28) | 16 (42.11) | 0 | 0 |
| Same job as before pandemic | 13 (19.40) | 13 (34.21) | 54 (91.53) | 31 (86.11) |
| Now working different job | 14 (20.90) | 3 (7.89) | 3 (5.08) | 2 (5.56) |
| Currently out of work<sup>2</sup> | 11 (16.42) | 6 (15.79) | 2 (3.39) | 3 (8.33) |
| Hours worked<sup>3</sup> |  |  |  |  |
| Fewer than usual | 16 (59.26) | 7 (43.75) | 17 (29.82) | 18 (54.55) |
| About the same as usual | 11 (40.74) | 7 (43.75) | 38 (66.67) | 15 (45.45) |
| More than usual | 0 | 2 (12.50) | 2 (3.51) | 0 |
| Employer COVID accommodations<sup>3</sup> |  |  |  |  |
| Made no accommodations | 13 (48.15) | 1 (6.25) | 46 (80.70) | 10 (30.30) |
| Made any accommodation | 14 (51.85) | 15 (93.75) | 11 (19.30) | 23 (69.70) |
| Provided masks<sup>4</sup> | 12 (85.71) | 14 (93.33) | 10 (90.91) | 21 (91.30) |
| Provided gloves<sup>4</sup> | 12 (85.71) | 14 (93.33) | 7 (63.64) | 18 (78.26) |
| Provided other PPE<sup>4</sup> | 2 (14.29) | 1 (6.67) | 3 (27.27) | 1 (4.35) |
| Provided hand sanitizer<sup>4</sup> | 11 (78.57) | 13 (86.67) | 5 (45.45) | 17 (73.91) |
| Cleaned surfaces more than usual<sup>4</sup> | 4 (28.57) | 11 (73.33) | 3 (27.27) | 7 (30.43) |
| Build partitions between workers<sup>4</sup> | 0 | 9 (60.00) | 0 | 6 (26.09) |
| Separated workstations<sup>4</sup> | 1 (7.14) | 6 (40.00) | 0 | 2 (8.70) |
| Conducting daily health checks<sup>4</sup> | 1 (7.14) | 7 (46.67) | 0 | 4 (17.39) |
| Closed break or eating spaces<sup>4</sup> | 1 (7.14) | 6 (40.00) | 0 | 3 (13.04) |
| Told to work from home | 0 | 0 | 0 | 1 (3.03) |
| Other<sup>4</sup> | 1 (7.14) | 1 (6.67) | 1 (9.09) | 1 (4.35) |

<sup>1</sup> 59 spouses reported for farmworker families and 36 for non-farmworker families
<sup>2</sup> Includes those who report having lost a job, being temporarily laid off, and on sick or other leave
<sup>3</sup> Based on number of individuals reported to be working
<sup>4</sup> Based on number of individuals who report employer made accommodations

Over half (59.29%) of the respondents from farmworker families reported working fewer hours now compared to prior to the pandemic; those from non-farmworker families were mixed, with only 43.75% reporting fewer hours and 12.50% reporting more hours. The difference between those in farmworker and non-farmworker families was not significant (p=.1628). For the spouses, most (66.67%) of those in farmworker families reported working about the same hours; a majority (54.55%) of those in non-farmworker families worker fewer hours. Differences for spouses were significant (p=.0430).

Significantly fewer respondents from farmworker families (51.85%) than non-farmworker families (93.75%) reported that their employers had made any accommodations for COVID-19 in the workplace (p=.0063). Among their spouses, only 19.30% among farmworker families reported accommodations, compared to 69.70% in non-farmworker families (p<.0001). The most frequent accommodations made in each group were providing masks, gloves, or hand sanitizer. Among workers from non-farmworker families, 60.00% of respondents and 26.09 of spouses reported that employers had built partitions between workers. Most of the other possible accommodations (e.g., conducting daily health checks, closing break or eating spaces, or telling employees to work from home) were reported by few workers; in all cases, more respondents from non-farmworker families reported accommodations for themselves and their spouses.

Concerns stated by women related to their own work differed in content and emphasis between the farmworker and non-farmworker family respondents (Table 3). Among those from farmworker families, the primary concerns had to do with the way their work was organized due to machinery and the way fields are laid out, and the difficulties presented in maintaining recommended physical distancing. Women were both planting and harvesting when interviewed in the late spring. Planting requires two or more workers sitting close to each other on the back of a mechanical planter pulled by a tractor and placing seedlings in revolving wheels that spaces them into the rows below the tractor. Harvesting blueberries, in the case mentioned, involves standing in narrow rows and working close to bushes, to be able to reach all around them to find the ripe berries. The other concern for women from farmworker families was problems in mask use. Comments suggested the women were not given proper masks, but used scarves to cover their face, which had to be removed due to heat.

**Table 3.**
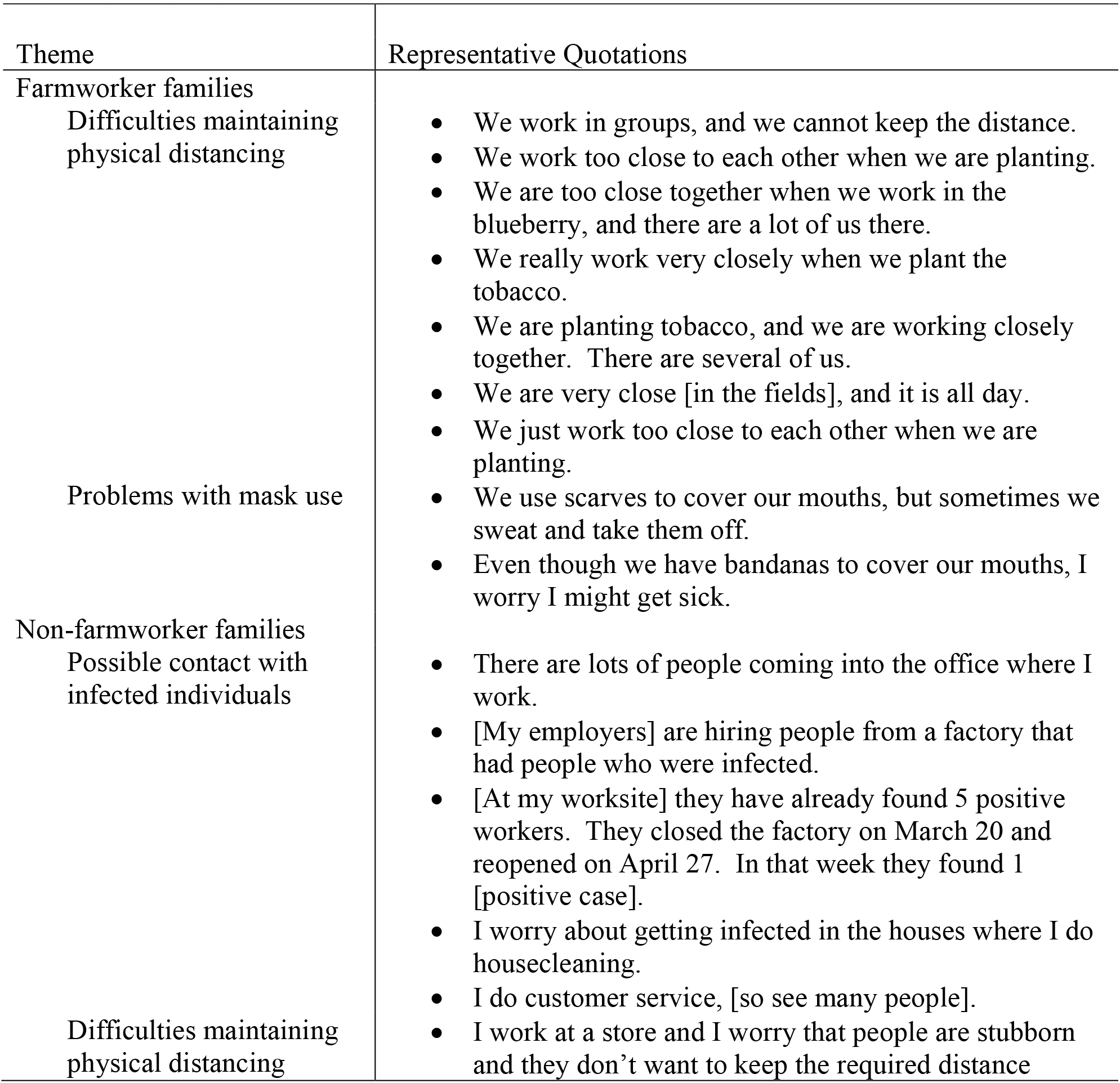
Concerns expressed by respondents about their own employment and risks of COVID-19, comparing respondents in farmworker and non-farmworker families.

For non-farmworker family respondents, the primary concern was coming into contact with infected individuals in the workplace. These women’s workplaces are more varied, so that is reflected in concerns ranging from infected individuals being hired into factories to cleaning houses where infected people may live, to encountering a variety of unknown people in offices and stores. Difficulties in maintain appropriate physical distance was mentioned, but less frequently than by women in farmworker families.

Concerns expressed by women for their spouses’ safety at work were more numerous, but were similar to concerns for themselves (Table 4). Women in farmworker families reported many concerns about mask use—that spouses did not use them in closed cars and while working. Most cited heat as a reason masks were not used. Physical distancing was also listed as a problem, particularly while doing farm work. There was also suspicion that co-workers might not be taking precautions and that that could lead to contact with infected individuals.

**Table 4.**
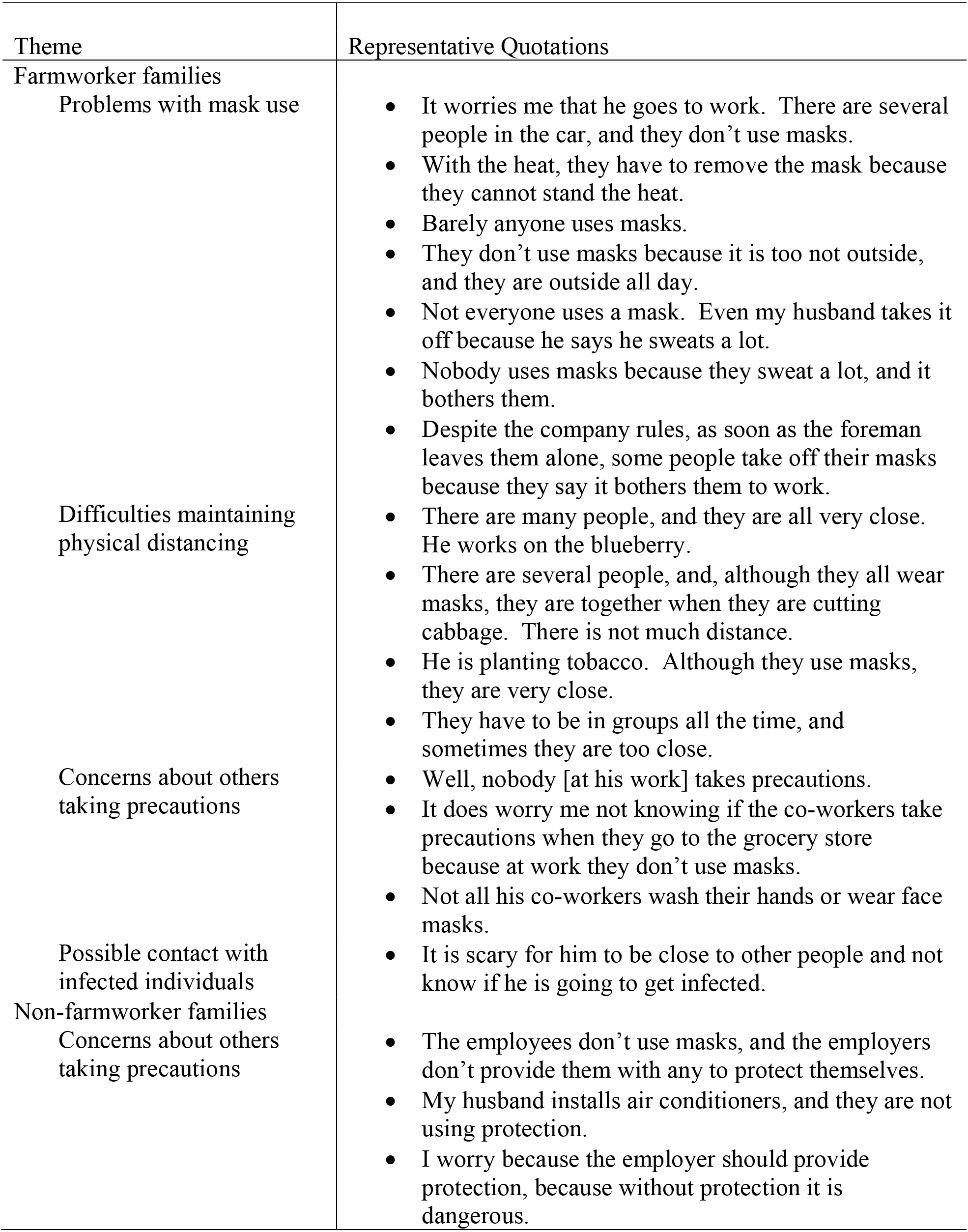
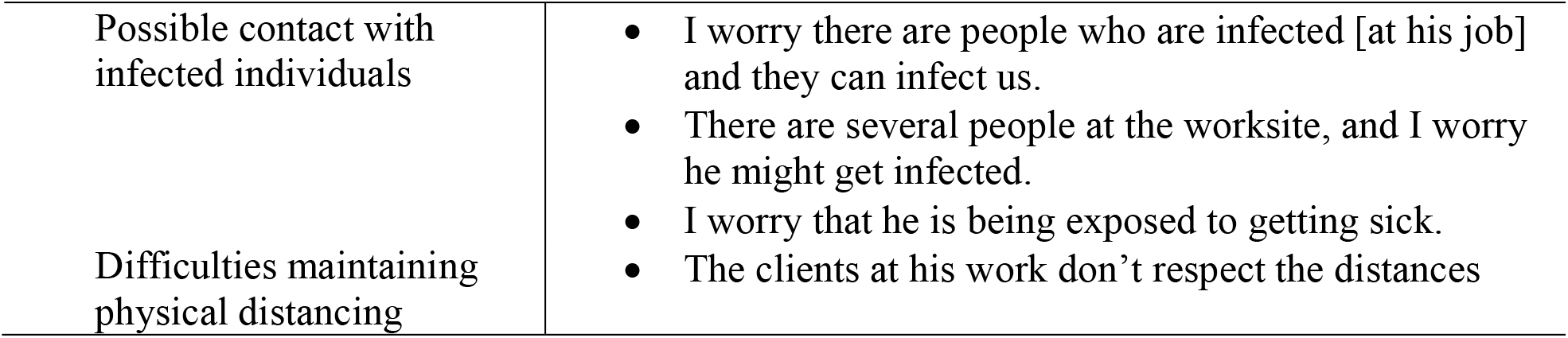
Concerns expressed by respondents about their spouse’s employment and risks of COVID-19, comparing respondents in farmworker and non-farmworker families.

Women in non-farmworker families had many of the same concerns. They were more direct than women in farmworker families about the possibility that their spouses could be infected and then bring infection home to the family.

#### Household economics

Most of both the farmworker and non-farmworker family respondents thought it was very likely or somewhat likely that they would run out of money for food in the coming three months (p+.0009) (Table 5). When current food security was compared, farmworker families reported significantly better food security than did non-farmworker families (p=.0062); 10.61% vs. 34.21%, respectively, reported some level of insecurity, with only non-farmworker families reporting actual behavioral measures taken to deal with shortage of food.

**Table 5.** Economic and food security concerns. Latinx farmworker and non-farmworker adults in North Carolina, May 2020.

|  | Farmworker<br>n=67 |  | Non-Farmworker<br>n=38 |  |
| --- | --- | --- | --- | --- |
|  | n | % | n | % |
| Likelihood family will run out of money in the next three months<sup>1</sup> |  |  |  |  |
| Very likely | 8 | 12.12 | 17 | 44.74 |
| Somewhat likely | 36 | 54.55 | 11 | 28.95 |
| Possible | 7 | 10.61 | 6 | 15.79 |
| Unlikely | 14 | 21.21 | 3 | 7.89 |
| Very unlikely | 1 | 1.52 | 1 | 2.63 |
| Highest level of food insecurity reported, past 7 days<sup>1</sup> |  |  |  |  |
| No food insecurity | 59 | 89.39 | 25 | 65.79 |
| Worried that food would run out because of lack of money | 7 | 10.61 | 10 | 26.32 |
| Anyone in the family ate less than they thought they should because of lack of money | 0 | 0 | 2 | 5.26 |
| Anyone in family go for a whole day because of lack of money | 0 | 0 | 1 | 2.63 |
| Conditions making it hard to get food family needs<sup>2</sup> |  |  |  |  |
| Grocery stores open fewer hours | 0 | 0 | 22 | 57.89 |
| Grocery stores make wait in line to enter | 0 | 0 | 20 | 52.63 |
| Grocery stores do not have food wanted | 0 | 0 | 18 | 47.37 |
| Children not getting school breakfast or lunch | 0 | 0 | 12 | 31.58 |
| Restaurants usually patronized are closed | 0 | 0 | 22 | 57.89 |
| Experiences no problems getting food | 67 | 100.00 | 4 | 10.53 |

<sup>1</sup> One missing farmworker observation.
<sup>2</sup> Excludes problems related to money.

Non-farmworker families cited numerous circumstances beyond money that made it difficult to get the food the family needed during the pandemic. Over half cited grocery stores being open fewer hours (57.89%) or making customers wait in line to enter the store (52.63%), as well as usual restaurants being closed (57.89%). About half (47.37%) noted that grocery stores did not have the food they needed, and a third (31.58%) noted that children not getting school breakfast or lunch made it difficult to meet family food needs.

#### Childcare and educational concerns

In both samples, most families accommodated childcare by having the parents work at different times (or a parent not work), so that one parent was always available (Table 6). As the second most used childcare option, almost a third (31.34%) of farmworker families reported leaving children home alone, while 21.05% of non-farmworker families reported that a friend or relative provided childcare. There were no significant differences in the use of different child care strategies, though twice as many farmworker families reported children staying home alone than non-farmworker families (p=.0797).

**Table 6.** Childcare and educational concerns. Latinx farmworker and nonfarmworker adults in North Carolina, May 2020.

|  | Farmworker<br>n=67 |  | Non-Farmworker<br>n=38 |  |
| --- | --- | --- | --- | --- |
|  | n | % | n | % |
| Child care while parents work<sup>1</sup> |  |  |  |  |
| Spouses work at different times | 38 | 56.72 | 26 | 68.42 |
| Leave children at home alone | 21 | 31.34 | 6 | 15.79 |
| Friend or relative provides childcare | 8 | 11.94 | 8 | 21.05 |
| Children attend licensed daycare center | 0 | 0 | 0 | 0 |
| Takes child to work | 1 | 1.49 | 0 | 0 |
| Education while school is closed |  |  |  |  |
| Received books or assignments | 67 | 100 | 36 | 94.74 |
| Received instruction over the internet | 67 | 100 | 38 | 100 |
| Concerns about children's education |  |  |  |  |
| Will fall below grade level | 64 | 95.52 | 37 | 97.37 |
| Will not graduate or be promoted | 23 | 34.33 | 20 | 52.63 |
| Will do poorly on end-of-grade tests | 10 | 14.93 | 22 | 57.89 |
| Cannot take college tests (e.g., SAT, ACT) | 1 | 1.49 | 7 | 18.42 |
| School closure have made parent work difficult |  |  |  |  |
| Disagree | 0 | 0 | 9 | 23.68 |
| Neutral | 45 | 67.16 | 13 | 34.21 |
| Agree | 22 | 32.84 | 16 | 42.11 |
| Satisfied with communication from schools |  |  |  |  |
| Disagree | 43 | 64.18 | 5 | 13.16 |
| Neutral | 22 | 32.84 | 5 | 13.16 |
| Agree | 2 | 2.99 | 28 | 73.68 |
| Children will be prepared for school next year |  |  |  |  |
| Disagree | 54 | 80.60 | 11 | 28.95 |
| Neutral | 13 | 19.40 | 9 | 23.68 |
| Agree | 0 | 0 | 18 | 47.37 |

<sup>1</sup> Respondents could report more than one childcare option.

All respondents for farmworker families reported that their children had received books or assignments when school was dismissed for the pandemic at the end of March. All but 2 of the non-farmworker families reported this, too. All respondents in both samples reported that at least some of their children received instruction over the internet while school was closed.

The concern that children would fall below grade level was the most frequently endorsed fear of parents, with over 95% in both groups expressing concern. Other concerns (that children would not graduate or be promoted, that children would do poorly on end-of-grade test, and that children could not take college tests) were endorsed more by non-farmworker families (p=.0980, p<.0001, and p=.0032, respectively).

Overall, respondents from farmworker families were less satisfied with school closure than those from non-farmworker families. They tended to agree that the school closure had made it more difficult to work or do household tasks (p<.001) and they tended to be less satisfied with communication from their children’s school to support learning (p<.001). They also tended to disagree more with the idea that their children would be prepared for school next year (p<.001).

#### Healthcare

Four (5.97%) farmworker family respondents reported that a family member had visited a doctor or clinic in the previous two weeks. None had canceled an appointment for any reason. In contrast, 11 (28.95%) of non-farmworker family respondent reported a doctor or clinic visit in the past two weeks. An additional 9 reported having had an appointment, but canceled it due to worries about the coronavirus. One of these respondents reported that she was afraid of getting infected and then infecting her family, if she went to the clinic.

#### Community attitudes

When asked about their community members’ current worries about racism and discrimination, there were sharp contrasts between farmworker and non-farmworker family responses (Table 7). For farmworker families, most (91.04%) responded that they did not know if community members were more worried. In contrast, non-farmworker families gave split responses: 42.11% reported more worry about racism and discrimination, while 57.68% reported no more worry than previously. Similarly, when asked about worry concerning immigration issues, most (89.55%) farmworker families responded that they did not know, and the rest said no. Only 21.05% of non-farmworker families thought there had been an increase in worry about immigration issues, while 78.95% said no.

**Table 7.** Concerns about community climate. Respondents were asked if they thought community members were more worried than usual about topics. Latinx farmworker and nonfarmworker adults in North Carolina, May 2020.

|  | Farmworker<br>n=67 |  | Non-Farmworker<br>n=38 |  |
| --- | --- | --- | --- | --- |
|  | n | % | n | % |
| Discrimination and racism |  |  |  |  |
| Yes | 2 | 2.99 | 16 | 42.11 |
| No | 4 | 5.97 | 22 | 57.89 |
| Don't know | 61 | 91.04 | 0 | - |
| Immigration issues |  |  |  |  |
| Yes | 0 | - | 8 | 21.05 |
| No | 7 | 10.45 | 30 | 78.95 |
| Don't know | 60 | 89.55 | 0 | - |

Farmworker families had few further comments on community climate, In contrast, non-farmworker families did express concerns. They noted that, as non-citizens, they were not being provided with financial assistance that citizens were receiving. One woman noted, “There are many people who have complained that we are not cared for the same [as citizens], and the language barrier does not help. We lose at everything. The government does not help with the relief from the President, who discriminates against those of us [even though we] pay taxes.” Priority for assistance for citizens was noted by women in financial matters, particularly in contrast for those without documents. Some believed that if they get sick, priority will be given to Americans in medical care. Another noted a specific example, that the pharmacy has started to be reluctant to give her child’s medicine to her without a US passport or US ID. Some perceived increased racism in their interactions with non-Latinx individuals. “People stare at us a lot in the grocery store,” one woman said. Another noted, “If you are in the US and Hispanic, they always look at you different; they look down on you.” An attitude to blame them for illness was noted: “I have heard people saying that Mexicans are going to get sick because they don’t take care of themselves.” Another noted, “They think that, because we are not from here, we are the ones that bring the diseases.”

Women in non-farmworker families also expressed fears related to immigration status that continue or are exacerbated by the pandemic. One stated, “If the President does not stop doing things, one is always at risk. There are many people deported, and people are still scared in this pandemic.” Another worried, “I am scared that, if we go out, they can detain me for lack of documentation.”.

#### Overall concerns

Fear, worry, and anxiety characterized the concerns expressed by women in rural farmworker families (Table 8). They were afraid of contracting COVID-19 and worried about bringing the disease home to their families. They seemed acutely aware that contracting COVID-19 might leave them unable to care for their children. Some had stopped work to care for their children, often because the cost of childcare (necessary because schools had closed) was more than they could afford with the low wages they earned. Economic concerns added to their worries, particularly paying for basic needs such as food and, if needed, healthcare for COVID-19. They were concerned for high-risk family members, and many knew someone who had been sick or died from COVID-19. Despite their concerns, they stated that they did not use masks when visiting households of relatives, either because they assumed that “family” would not be a source of infection or because they thought it would not “look good” to wear masks around relatives. One woman’s comment seemed to sum up those of the group: “This whole situation is stressing me out and giving me anxiety.”

**Table 8.**
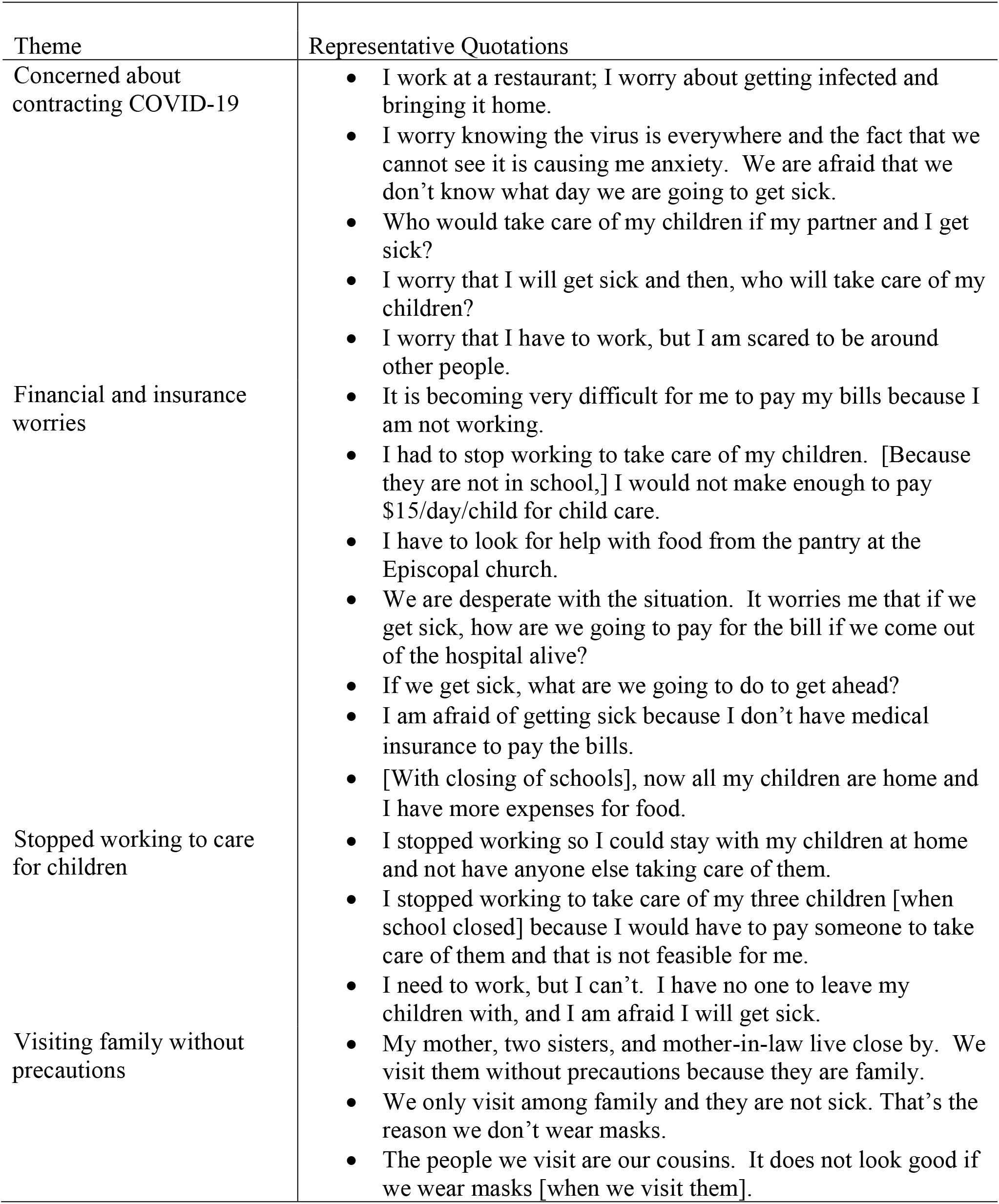
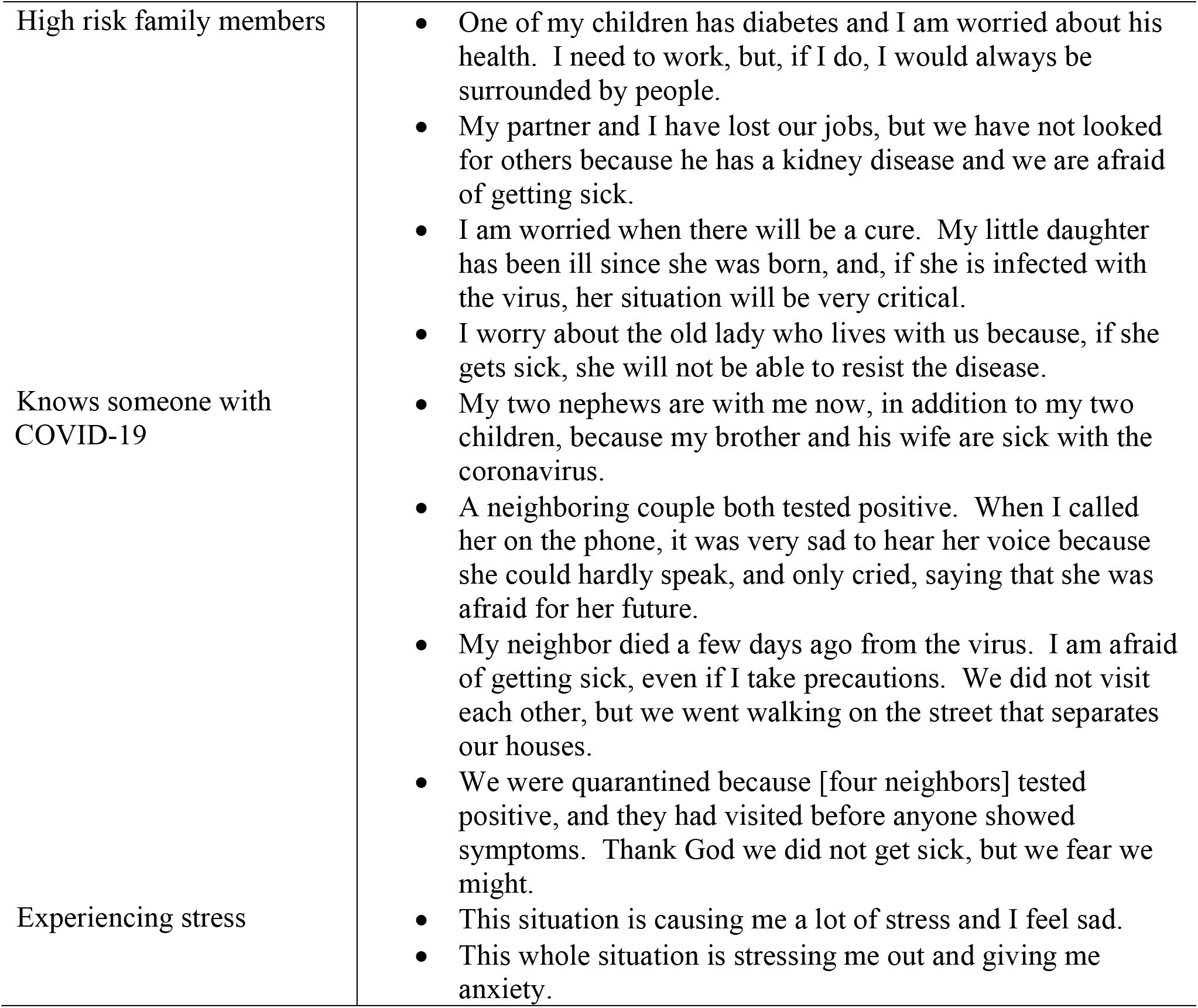
Comments by respondents from farmworker families about their own situation during the COVID-19 pandemic.

Women in urban non-farmworker families had concerns that reflected the lower rates of employment in this group (Table 9). Like the women in rural farmworker families, worry and fear characterized their responses. They also had a different worry about their children that was not expressed by the rural women, that children were at home and could not go outside.

**Table 9.** Comments by respondents from non-farmworker families about their own situation during the COVID-19 pandemic.

| Theme | Representative Quotations |
| --- | --- |
| Worries about children | • I am worried that, because of the pandemic, the children are not able to go out.<br>• This whole situation worries me and especially that the children can't go out.<br>• I am very concerned about my children because [the current situation] is not that same as when they go to classes and are taught by their teacher. It is very different at home.<br>• I am concerned about the children. They are not made to be locked up, though it is for their safety that they cannot out. |
| Concerns about contracting COVID-19 | • People don't understand that they need to protect themselves by wearing masks and staying home.<br>• I am worried that my daughters will get infected.<br>• We are scared of getting sick.<br>• Everyone is afraid of getting [COVID-19]. Some people are not protecting themselves. |
| Financial and insurance worries | • I am worried because I don't have work, and it is worse in my country.<br>• I am worried for my family because my husband does not have full time work.<br>• We don't have work, so we need to be directed to places that help pay rent. |
| Experiencing stress | • I am worried because something else new comes out every day, and the numbers [of cases] are increasing.<br>• I am worried about everything. |
| Knows someone with COVID-19 | • We had a visitor with a fever who tested positive, and he had contact with my children |

## Discussion

This survey of rural farmworker families and urban non-farmworker families demonstrated that these immigrants face a number of impacts of COVID-19 and that these impacts are sources of concern. About 1 in 4 women, both rural and urban, was out of work several months into the pandemic. Across both groups, many women and men were working fewer hours than before the pandemic. Rural women reported fewer worksite accommodations for safety in the COVID-19 pandemic for themselves and their spouses than did urban women. Across the entire sample, only one person (an urban spouse) had been told to work from home. These work-related issues translated into economic concerns. These were more severe among the urban non-farmworker families, among whom a much higher percentage viewed the likelihood of running out of money in the next three months as very likely. Similarly, urban women reported more food insecurity in the previous week and reported that restrictions in grocery stores and restaurant closures made it hard to get the food their families needed. Although almost all women reported children had received school books and assignments, most had concerns about how their children would learn at home. While urban women were generally satisfied with communications from the schools and thought children would be prepared or the next school year, rural women did not share these sentiments. Urban women expressed greater concern about community climate about racism and immigration issues than did rural women.

These findings highlight the diversity of experience Latinx families report during the pandemic, which likely reflect differences between the rural and urban environments as well as between the two populations. The rural health literature has consistently shown that rural populations have less access to quality medical care and greater age-adjusted mortality.^30^ Such statistics are often linked to lower household incomes, less formal education, as well as distance to both formal medical services (e.g., specialty care) and health-promotion services (e.g., healthy food distribution). Environmentally, rural communities have consistently lower levels of investment in the social determinants of health such as housing, emergency services, libraries, parks, and other services than urban communities.^31^ Socially, rural communities are often thought to provide stronger notions of community and support, based on a willingness to work together and pride in place.^30^ These rural-urban differences observed in other situations are reflected in this study. Even among these immigrants, the urban sample has higher levels of education. Almost 29% of urban women were at least high school graduates, versus 16% of the rural sample; among men the difference is almost 31% versus 9%. Workplace safety responses also appear better in the urban setting. About 48% of rural women and 81% of their spouses report no workplace COVID-19 accommodations, compared to only 6% of urban women and 30% of their spouses. The rural women had trouble answering questions about the impact community attitudes toward them as immigrants, while over 40% of urban women reported they were worrying more about racism and discrimination. As farmworkers are essential to rural agriculture and its economic contribution to the community, it is not surprising that there were fewer negative attitudes experienced.

Despite the differences between the rural and urban concerns, women in both samples expressed similar emotions and resulting mental health effects. Fear and worry were expressed by many women. They are afraid they will get sick and not be able to take care of their children. They worry that they have to go to work, but do not know if they will be exposed to COVID-19 by co-workers or others they encounter in the workplace. They are afraid they will not have enough money to pay bills. They worry about being exposed to the virus and bringing it home. They express desperation, anxiety, and stress in reaction to this fear and worry. Mental health data gathered nationally early in the pandemic show increased effects on mental health^32^ and suggest that the effects will result in more mental health impairment in disadvantaged groups, such as immigrants, as they cope with greater financial insecurity and grief caused by greater pandemic morbidity and mortality.^33^ In addition, we have shown elsewhere^10^ that these women are well informed about the causes of COVID-19 and recommended practices to prevent it. Their comments here reflect this knowledge and their inability to practice some of the methods to prevent COVID-19 in their families.

Understanding the impact of the COVID-19 pandemic on immigrants is aided by the use of a social justice framework and the idea of structural violence.^34-36^ Scholars have noted that exposure to social injustices results in effects over time on well-being, effects termed structural violence. Generally, such effects are slow and, some^34^ would argue, silent, reflecting the accumulation of exposure to risks over time, “slow violence”.^37^ However, events such as environmental disasters (e.g., hurricanes, wildfires, pandemics) place those suffering from slow violence at extreme risk, what has been termed “contextual vulnerability”.^38-40^ Both the long term compromised well-being and the socioeconomic context in which the disaster occurs focus risk on the immigrant Latinx population. In the current study, this structural vulnerability played out in several ways, but economic effects are most evident. Because access to resources and the ability to reduce exposure and recover from the effects of the pandemic are not uniformly distributed, Latinx immigrants are forced to (in the case of women) reduce household income by giving up low paying jobs to care for children receiving school instruction at home, while spouses continue to experience disease exposure in their own low paying jobs that are either deemed essential to society or are, in fact, essential to their household’s survival. While the essential nature of some jobs, e.g., “first responders”, has been met with extra benefits from public demonstrations of support to largescale donations of free goods and services to designated grocery shopping hours. Few of these benefits filter down to immigrant workers whose work is often as structurally essential to the work of first responders as it is to society at large.

Comparable rewards (like free meals) that support healthcare workers during the pandemic are also needed and deserved by the very people doing the work to bring this food to the tables across the nation. Unfortunately, rather than seeing these benefits, immigrants are actually suffering from food insecurity.

This study has several limitations that should be considered. All behaviors were self-reported and not observed. The women interviewed also reported for their spouses. Data could not be verified and may be inaccurate or distorted because of social desirability. Small sample sizes prevent more detailed analyses of quantitative data. There was greater uniformity in occupations within the rural group, which makes their responses easier to interpret.

Nevertheless, this study has several strengths. One was the scheduling of data collection within a short time (the month of May) during which changes in national information about prevention and state regulations were relatively stable. By the time of data collection, emerging research had established the importance of physical distancing and mask use over the initial emphasis on hand hygiene and cleaning surfaces.^41-43^ Another strength was that all families in this study would have been subject to the same North Carolina governmental orders.

## Conclusions

Overall, this study represents a unique opportunity to document the concerns of Latinx immigrants in the USA during the early days of the COVID-19 pandemic. In particular, farmworkers are often a hidden and difficult to reach population. This study demonstrates that both rural and urban Latinx immigrants have substantial concerns about their families’ vulnerability in the COVID-19 pandemic. While the content of these worries differs some between urban and rural, the emotional response is the same. The information provided by the women in this study indicates that concerns are well founded. Workplaces are not providing protections for workers, and the need to provide childcare has taken many women out of the labor force. At least in the urban areas, families are encountering more overt racism and are worried about immigration issues.

The results of this study illustrate the contextual vulnerability of Latinx immigrants in North Carolina. Several policy reforms can help to alleviate this. First, immigration reform is needed so that those immigrants doing essential work can access benefits available to other workers, including sick leave, family leave, nutrition benefits, and healthcare. Second, work safety culture and work organization need to be addressed so that workplaces are safer for all illnesses and injuries, including infectious disease. Third, access to education and job training benefits need to be made equitable so that the jobs held by this population can be higher paying and so their children can make better use of school provided resources.

## Data Availability

The data are not currently available.

## Acknowledgments

This research was funded by a grant from the National Institute of Environmental Health Science, Grant Number R01 ES08739.

The authors appreciate the support and participation of their community partner, the NC Farmworkers Project, and of Student Action with Farmworkers. They also appreciate the valuable contributions of our community field interviewers in carrying out participant recruitment and data collection. They especially thank the mothers who participated in this study.

